# Homegrown Ultraviolet Germicidal Irradiation for Hospital-Based N95 Decontamination during the COVID-19 Pandemic

**DOI:** 10.1101/2020.04.29.20085456

**Authors:** Eric Schnell, Melanie J. Harriff, Jane E. Yates, Elham Karamooz, Christopher D. Pfeiffer, James F. McCarthy, Christopher L. Trapp, Sara K. Frazier, John E. Dodier, Stephen M. Smith

## Abstract

Coronavirus disease (COVID-19), the disease caused by the severe acute respiratory syndrome coronavirus 2 (SARS-CoV-2) virus, is responsible for the 2020 global pandemic and characterized by high transmissibility and morbidity. Healthcare workers (HCWs) are at risk of contracting COVID-19, and this risk is mitigated through the use of personal protective equipment such as N95 Filtering Facepiece Respirators (FFRs). The high demand for FFRs is not currently met by global supply chains, potentially placing HCWs at increased exposure risk. Effective FFR decontamination modalities exist, which could maintain respiratory protection for HCWs in the midst of the current pandemic, through the decontamination and re-use of FFRs. Here, we present a locally-implemented ultraviolet-C germicidal irradiation (UVGI)-based FFR decontamination pathway, utilizing a home-built UVGI array assembled entirely with previously existing components available at our institution. We provide recommendations on the construction of similar systems, as well as guidance and strategies towards successful institutional implementation of FFR decontamination.

## Background and Rationale for Ultraviolet-C Germicidal Irradiation

One challenge that has emerged during the current COVID-19 pandemic is that the SARS-CoV-2 virus appears to frequently infect health care workers, threatening to deplete the pool of people available to care for sick patients at a time when they are most needed. Respiratory protection is an important component of interrupting the chain of SARS-CoV-2 transmission. The Centers for Disease Control and Prevention (CDC) currently recommends N95 Filtering Facepiece Respirators (FFRs) or equivalent, when available, for care of COVID-19 positive patients [1]. Unfortunately, N95 FFRs are currently in critically short supply due to the intersection of dramatically increased global demand and fractured supply chains. Thus, an urgent need exists for the development of decontamination strategies for used N95 FFRs during this crisis.

N95 FFRs can be decontaminated and re-used several times without loss of protective efficacy [2–9]. Although a wide variety of decontamination modalities effectively decontaminate N95 FFRs, some of these modalities (such as spraying with alcohol) degrade the mask material’s aerosol filtration abilities or alter subsequent FFR fit [4]. Other modalities, such as vaporized hydrogen peroxide, are highly effective and preserve FFR function [4], but require equipment that could be difficult to obtain and operationalize during a pandemic.

One effective modality for N95 FFR decontamination involves ultraviolet-C germicidal irradiation (UVGI). After the 2003 severe acute respiratory syndrome (SARS) epidemic, numerous studies demonstrated that coronaviruses, similar to SARS-CoV-2, are inactivated by light in the UV-C spectrum (200–280nm) [6]. Inactivation occurs by photochemical degradation of the genetic material of coronaviruses, which is composed of single-stranded RNA (ssRNA) [10,11]. Institutions have developed and disseminated streamlined protocols for UVGI-based N95 FFR decontamination [10], on which our approach was initially based. However, similar to the challenges in acquisition of specialized equipment to implement other decontamination methodologies (e.g., vaporized hydrogen peroxide), commercial UVGI systems have also been difficult or costly to obtain during the current COVID-19 pandemic. In this document, we discuss technical considerations, practical guidance, and alternative approaches to the development of local facilities and methodologies for UVGI-based N95 FFR decontamination.

## Choice of UVGI Exposure Dose

The two critical aspects that must be considered by any N95 mask decontamination program are decontamination efficacy and subsequent respirator performance. An optimal dose of UV-C light must reduce pathogens on the N95 by at least 3-log levels (per recent FDA guidance [11]), without degrading mask function or fit.

On a flat, nonporous surface, UV-C exposure (254 nm peak wavelength) at a total energy “dose” of 2.5 - 6.5 mJ/cm^2^ reduces ssRNA virus levels by 2-log levels (99%), with a > 5-log reduction of virus at 20 mJ/cm^2^ [6, 12]. However, N95 FFRs are composed of multiple layers of material which are not fully penetrable by that UV-C dose, and contain uneven surfaces that UV-C light may not reach effectively due to shadowing. Thus, the efficacy of UV-C light in killing viruses located on mask material was evaluated independently. The research and engineering firm Applied Research Associates (ARA) performed UV-C validation studies on N95 FFRs using influenza H1N1 virus which, like SARS-CoV-2, is an ssRNA enveloped virus. They demonstrated that the UV-C dose required to reduce H1N1 virus to undetectable levels on N95 FFRs was 1 J/cm^2^ [13]. In a separate analysis, UV-C intensity of 1.0-1.2 J/cm^2^ also effectively decontaminated a wide variety of N95 FFRs (shapes/models) of H1N1 virus, even when viral inoculations were covered with soiling agents (artificial skin oil or artificial saliva) [5]. However, mask straps were more variably decontaminated at that dose, with several models achieving less than a 3-log reduction in viral load [5, 13], suggesting that an alternative modality, such as disinfectant wipes, might need to be applied to decontaminate straps of certain mask models.

UV-C light is well-validated to provide effective decontamination without degrading mask performance. UV-C exposure at doses up to 950 J/cm^2^ did not increase particle penetration beyond the 5% threshold for a variety of N95 FFR models tested in a prior analysis [2], and other physical properties of the masks, such as mask strap strength, remained functionally intact at doses up to 2500 J/cm^2^ for various FFRs [2]. Although mask filtration performance is not significantly affected by UV exposure, multiple cycles of use and decontamination (>10) decrease strap performance in some models [13]. However, multiple N95 don/doff cycles alone (without decontamination) are thought to adversely affect mask fit and strap performance [14], which are could become an issue before degradation from UV-C light reduces performance.

Based on the above analysis, we chose 1.0 J/cm^2^ as a minimum UV-C dose for mask decontamination, which is also consistent with recently released guidance from governmental and non-governmental agencies [15, 16]. The UV-C unit used at the VA Portland Health Care System delivers 1.0 - 2.0 J /cm^2^ to each N95 FFR, depending on the location of each component of a given N95 FFR within the UV-C exposure unit.

## UVGI Chamber Design and Construction

Light sources capable of producing UV-C light are widely commercially available when supply chains are functioning. During the COVID-19 pandemic, many “implementation ready”, commercial UVGI systems became unavailable, so we designed and constructed an in-house UVGI system, using equipment already available on site. Apart from biological safety hoods, UV-C bulbs are used in HVAC and water decontamination applications, and are frequently available from a hospital’s physical plant. A variety of bulbs exist for this purpose, which can vary both in power as well in terms of far UV-C wavelength production (175–210 nm). Far UV-C light can generate hazardous ozone gas [17], and additional design considerations would be necessary if these wavelengths were produced.

We designed and built a chamber specifically for UVGI decontamination of N95 FFRs using a local supply of low-pressure UV-C bulbs (Philips TUV G30T8 30W / 35 inch bulbs; dominant emission at 254 nm, minimal emission at 175–210 nm). Our design consists of two planar arrays of UV-C bulbs facing each other, producing a strong and uniform field of UV illumination to both sides of each mask and allow for simultaneous decontamination of multiple masks (**Figure 1**). Our electrical shop assembled our arrays using standard light mounts (‘tombstones’) and regulated power supplies (ballasts; see **Table 1** for materials). Each array is 4 bulbs wide (12 cm apart) and 3 bulbs long end-to-end (12 bulbs per array), and the distance between arrays was 70cm (**Figure 2A**). FFRs are suspended from stretches of nylon line (Black and Decker), strung between two hooks with a thick rubber band at one end to maintain tension.

**Figure 1.**
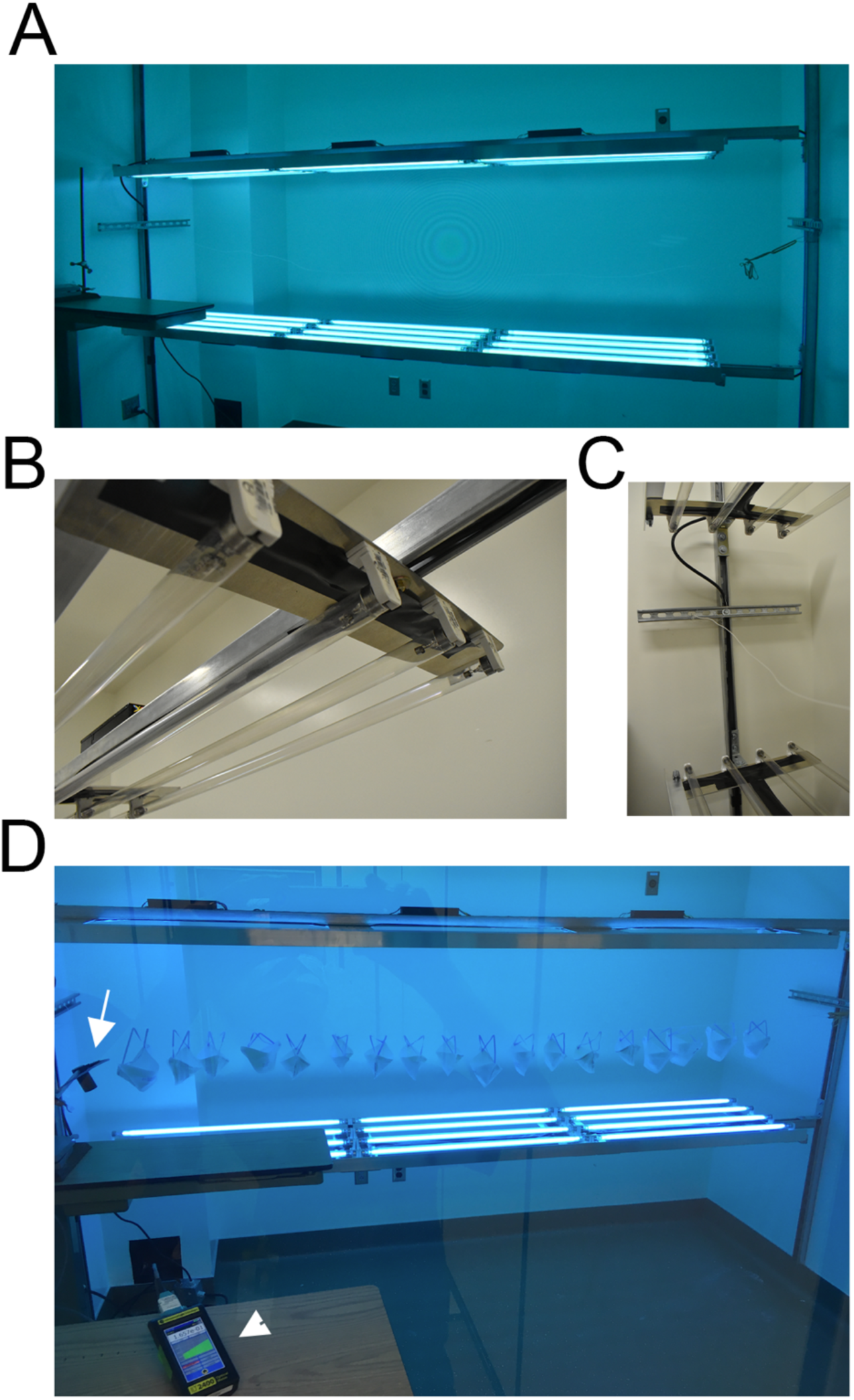
UVGI Arrays. A. Each array consists of 12 UV-C bulbs in a 3×4 array, facing each other. B. Longitudinal end of one array showing lampholder mounts on aluminum stock. C. Arrays mounted to Unistruts, with crossbar for lines to hang N95s. D. Masks arranged on a single line between UV-C arrays; light dosimeter probe on left (white arrow) mounted to measure light at iso-irradiant site relative to lowest exposure of each mask. Light meter console (arrowhead) in foreground for monitoring total dose delivered (as seen from clean workroom).

**Table 1.**
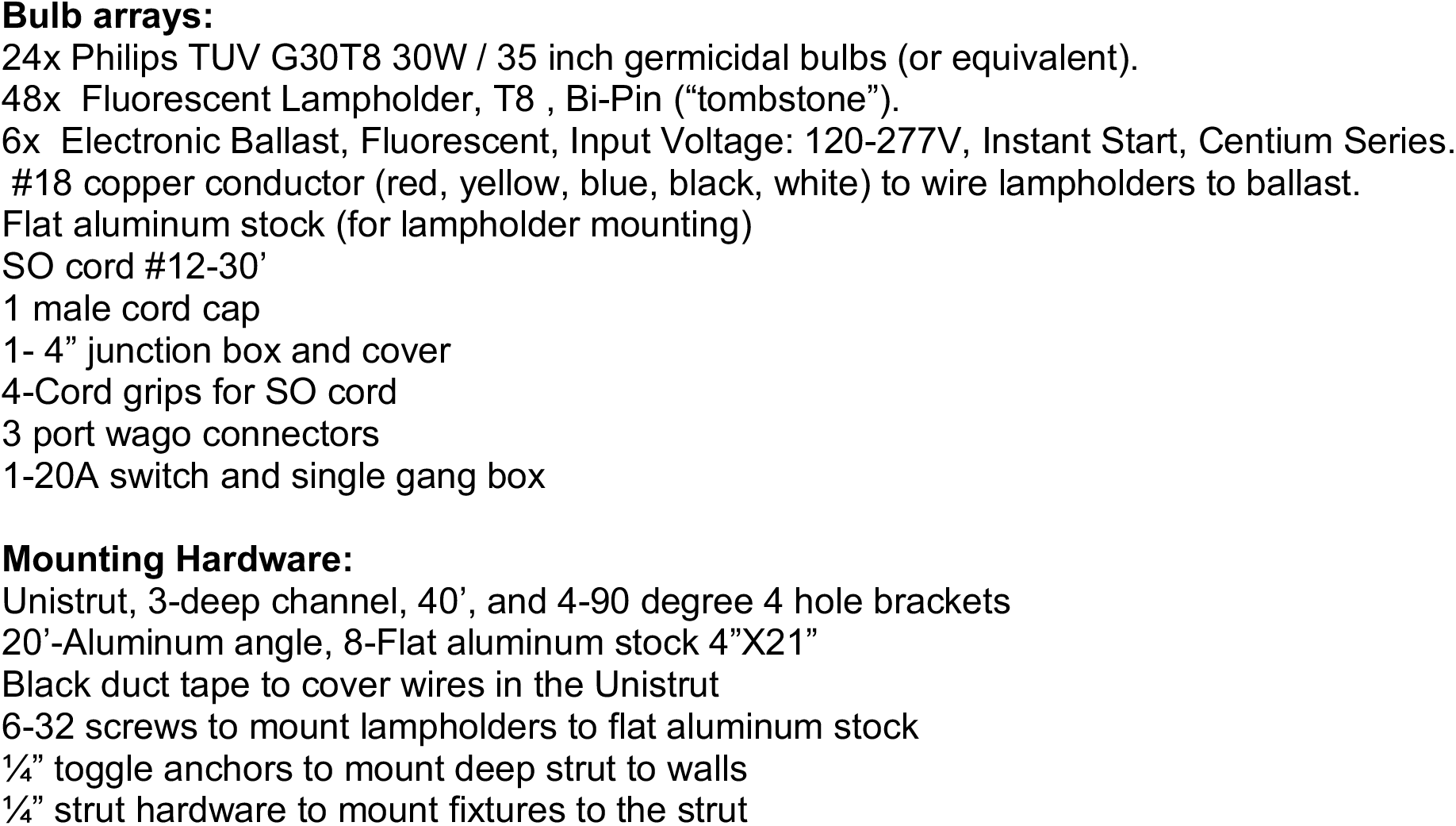
Construction Materials for UV-C bulb arrays.

Irradiance diminishes as one moves away from a light source, as described by the inverse square law. This relationship means that shorter inter-array distances provide stronger irradiance (and possibly greater rates of decontamination and throughput), but result in a greater difference in irradiance between the proximal and distal aspects of each FFR facing each array. Our intent was to provide a strong irradiance with only moderate variation over the region occupied by the N95s directly between the arrays. Between 28–42 cm from each array (the space occupied by the hanging masks directly between the arrays), our arrangement produces UV-C irradiance of 0.40-0.70 mW cm^2^ (**Figure 2A-C**). This range corresponds to the closest and furthest FFR components facing each array, as shown in **Figure 2C**. This irradiance provides a dosage of 24–42 mJ/cm^2^ in one minute, and 1.0-1.4 J/cm^2^ in approximately 30 minutes. Irradiance was observed to drop off markedly in the outermost 10 cm of the bulb array (**Figure 2B-C**), and these regions are not used for decontamination.

**Figure 2.**
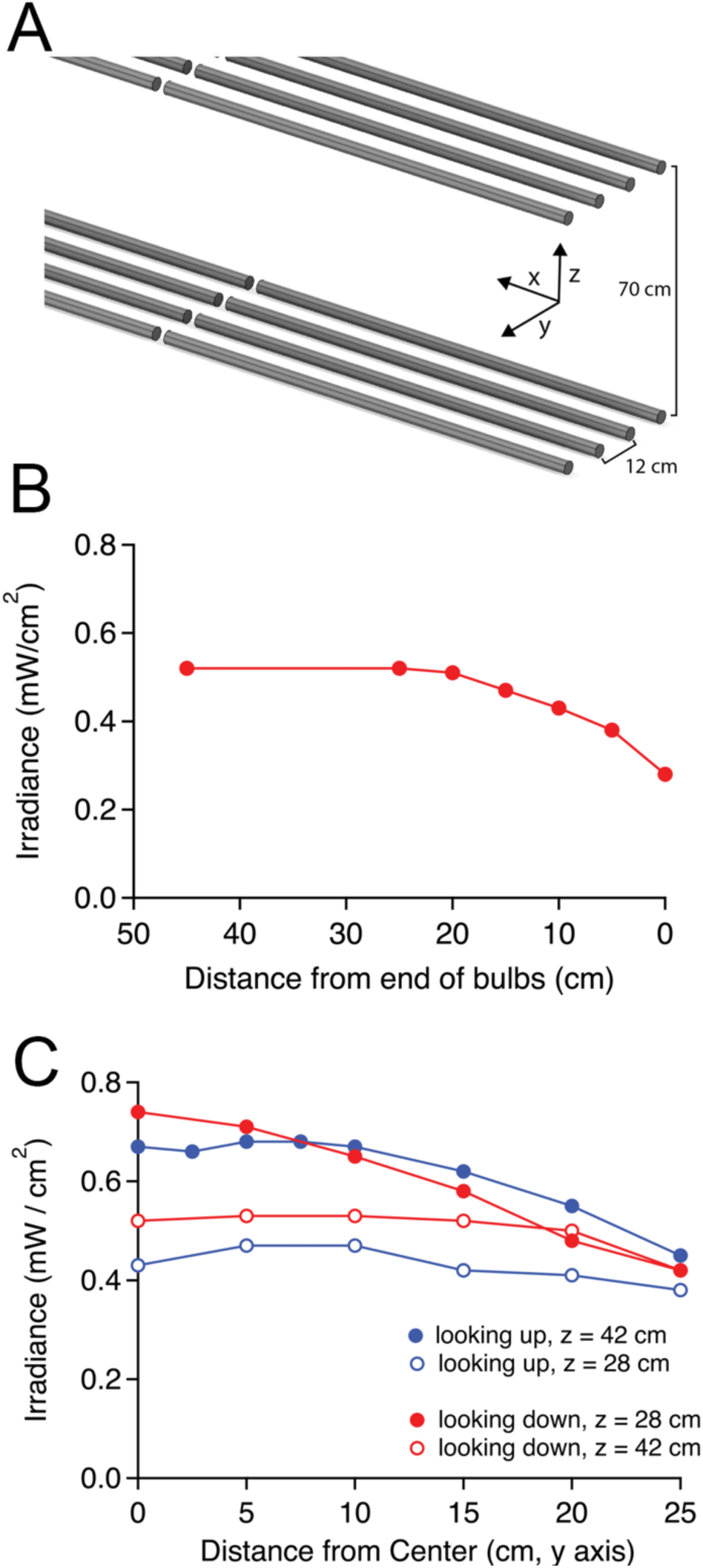
UV-C light intensity measured across each array. A. Schematic demonstrating array dimensions and axes. B. Light intensity variation along longitudinal stretch of each array beginning at the terminal tombstone ( = 0 cm) moving inwards along the x axis between the center bulbs. C. Light intensity variation moving outwards from the center of the linear array (along the y axis). Measurements recorded with dosimeter facing upwards/downwards at furthest vertical extent of each hanging FFR (in this case, z = 28 and 42 cm from bottom array, showing ranges of light irradiance).

Light fixtures were mounted onto flat aluminum stock bases, with mounting bars bracketed across a 8×10 ft room and bolted to walls (**Figure 1**). An additional safety rail was constructed on the staff side of the array to protect bulbs and staff from inadvertent contact. UV-C exposure is harmful to skin and eyes, so UV-C-blocking barrier (glass) was set up as an engineering control to protect staff from UV-C exposure during decontamination, while still allowing UVGI technicians to observe the light meter readings. UV-C light did not penetrate the glass, as verified by our UV-C dosimeter.

## UVGI Intensity Calibration and Measurement

One critical aspect in any UVGI-based decontamination process involves determining and controlling the total UV-C exposure delivered (“dose”) to ensure adequate N95 FFR decontamination in regards to previously validated exposure energies. Irradiance can vary significantly between bulbs, and even for individual bulbs varies over time, and thus we strongly recommend acquisition of a UV-C specific light meter when possible.

To measure UV-C dose, we used the ILT2400-UVGI meter (International Light Technologies, Peabody, MA), which is selectively sensitive to the wavelengths in the UV-C spectrum and reports UV-C light irradiance in W/cm^2^. This dosimeter can also integrate light exposure in real time, and report total UV-C dose delivered in J/cm^2^. We mounted this detector inside of our UVGI chamber to verify effective light delivery with each UVGI decontamination cycle, and have positioned it to measure light intensity in a direction/location corresponding to the least amount of UV-C energy that would be received by any FFR component (see below), thus ensuring that all mask components receive at least the minimum desired energy level. An optimal UV-C dosimeter must be accurately calibrated, which can be verified through ISO17025 accreditation (quality assurance) and NIST traceability (calibrated to a known source), and ideally register both irradiance and total energy delivered for ease-of-use.

In the absence of a specific UV-C dosimeter to this purpose, an alternative strategy could involve re-purposing other available research equipment to measure UV-C wavelengths. In our earliest implementation, we initially used a PM160T thermal power detector (ThorLabs, Newton, NJ), which detects wavelengths of 190 nm-10 μm, as this was immediately available in our laboratory for calibration of light intensities for optogenetics experiments [18]. The meter provides readings in mW units, which are converted to mW/cm^2^ by dividing measurements by detector surface area (0.785 cm^2^), and to mJ/cm^2^ by multiplying by exposure time in seconds. This small handheld meter was best utilized together with the manufacturer’s software to display power in graphical format, to zero the detector remotely via a long USB cable, and time average the readings. Time averaging also helped compensate for the sensitivity of this detector to other wavelengths, which caused readings to fluctuate near infrared sources (e.g., a gloved hand) or other field-generating electronics. This second issue could potentially be addressed by adding a shortpass optical filter to the detector array. Other home-designed UVGI measurement solutions utilizing photodiodes have also been evaluated [19].

## Alternative Chamber Designs

Prior to constructing our UVGI chamber, we implemented two additional UVGI N95 decontamination sites which we will describe here as they might be more quickly attainable. Both involved re-purposing of standard cell culture biosafety hoods, which are widely available in facilities with biomedical research labs and have been also been previously evaluated for N95 FFR decontamination [19].

First, our unmodified biosafety hood (SterilGARD, The Baker Company, Sanford, ME) contains a single UV-C bulb in its stock configuration, as well as a UV-C impermeable sliding glass door to protect staff. The bulb in our hood was a year-old GE G64T5 65W bulb, with an expected light intensity of 0.26 mW/cm^2^ at 1 m distance. We measured an irradiance of 0.18 mW/cm^2^ of UV-C intensity at the working surface directly below the bulb (approximately 60 cm away from the bulb) and 0.13 mW/cm^2^ at the working surface at the front of the hood (approximately 75 cm away from the bulb). A raised platform one foot from the center of the bulb received 0.34 mW/cm^2^. For comparison, the ClorDiSys Torch units implemented in Nebraska Medicine’s N95 decontamination suite produce 0.20 mW/cm^2^ at a distance of 10 feet [10, 20]. The main advantage of utilizing an already available biosafety hood is the potential for immediate implementation, as these units are widely available in biomedical research and clinical laboratories. However, one disadvantage of these units is that N95 FFRs can only be irradiated from one direction at a time, and thus require multiple exposures to decontaminate all mask surfaces. This would need to be performed carefully to avoid spreading potential contaminant from non-exposed to exposed surfaces. A second disadvantage is the time required to reach the necessary 1 J/cm^2^ dose for each surface (over 4 hours per mask with two exposures at 0.13 mW/cm^2^) as well as the 2-fold variability in UV-C irradiance at different lateral locations within the hood [19].

With minor modifications, we were able to significantly improve the workflow and light intensity inside the UV hood. We placed two light fixtures into the hood, at the top and bottom surfaces, to allow for simultaneous UVGI decontamination of both surfaces of each N95 mask. These fixtures were built using standard fluorescent fixtures (tombstones and ballasts) otherwise similar to the custom-built UVGI suite described above, with Philips TUV G30T8 30W and shorter Philips TUV G25T8 25W bulbs (also already on site). The upper fixture was suspended from the top of the box by chains, which resulted in a separation of 48 cm between the two bulb arrays (**Figure 3**). The outlets inside the hood are operated from a switch outside of the box, and thus could be operated remotely when the safety door was closed. The intensity of this light at the vertical midpoint of the chamber was 0.66 mW/cm^2^ facing either direction, but was uniform moving laterally until the last 25 cm of each bulb run. Thus, depending on the particular bulbs available at any individual institution, biosafety hoods could be safely modified to accommodate additional light sources to provide bidirectional irradiation.

**Figure 3.**
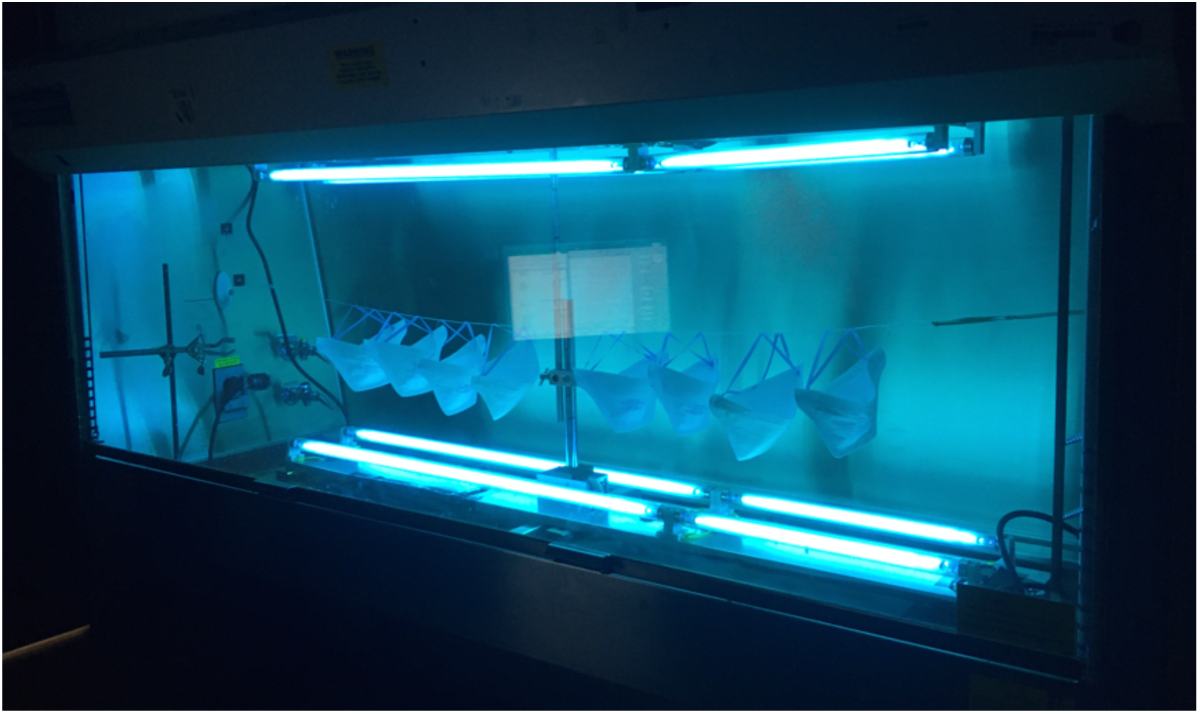
UVGI decontamination chamber constructed within a modified cell culture biosafety hood. Two identical four-bulb UV-C arrays were placed facing each other within the hood, and were used to decontaminate masks prior to construction of the larger array (Figure 1).

## Limitations and risks of UV-C decontamination

Although UV-C-based N95 FFR decontamination is effective and can be achieved with a variety of either commercially available or custom-built chamber designs, it does have several limitations and risks. These include the potential requirement for a secondary decontamination method for the straps on certain N95 FFR models [5, 13], and the observation that more resistant pathogens, such as bacterial spores, might not be inactivated with a 1 J/cm^2^ dose [13, 15]. Additionally, other FFR models, including non-surgical N95s or internationally-sourced KN95 FFRs, have not been formally evaluated in terms of their penetration to UV-C light, and thus might be variably decontaminated, as UV-C transmission through N95 layers is material-dependent [21]. Some UV-C bulbs also produce ozone, which is hazardous and should be appropriately vented.

Finally, we noticed a nutty/smoky odor when masks were donned shortly after UV-C decontamination. This did not affect fit-testing and was noticed to “off gas” spontaneously after several hours. Although this odor has not been fully characterized, a limited analysis performed by Applied Research Associates demonstrated that levels of 62 volatile organic compounds were either undetectable or several orders of magnitude below permissible exposure limits [22]. We recommend that off-gassing time be considered in UVGI FFR decontamination protocols when feasible.

## Process Implementation

The evaluation, construction and successful implementation of an N95 decontamination process (by UVGI or other methods) requires participation and involvement by multiple stakeholders throughout the hospital. One significant requirement is that decontamination efforts are set as a priority by facility leadership. Leadership priorities could empower rapid actions toward decontamination, primarily through the commitment of both staff and resources towards the effort, and with an agreement to address any needs as they arise for the effort. Once decontamination has been approved and prioritized from above, the highest priorities would be: 1 Formation of a workgroup to determine modality and feasibility (see below), 2. Assessment of available local resources that could be allocated towards decontamination efforts (for example, UV-C light sources and light meters), 3. Attempts to acquire necessary materials, as these are likely to be amongst the earliest rate-limiting steps, and 4. Determination of institutional regulatory/competency requirements, so that any appropriate forms and SOPs can be approved by required system entities, ideally in an expedited manner. Ideally these could be accomplished by workgroup members in parallel. Finally, FFR collection should begin as soon as possible, to preserve supply and to provide masks for decontamination once available.

## Workgroup: Composition and Integration

We recommend the formation of a workgroup with delegates from front-line healthcare providers, nursing management, sterile processing services, infection control, occupational safety, research staff, facilities staff and electricians, supply chain management, and facility leadership. Each of these groups will play a critical role in either engaging with, or facilitating, decontamination efforts. Possible roles for each group in the effort:

**Healthcare providers:** use and labeling of N95 FFRs, commitment to process
**Nursing Management:** implementation, coordination, communication
**Sterile Processing:** collection, tracking, regulatory compliance, SOP generation
**Infection Control:** decisions re: deployment of decontaminated N95s,
**Occupational Safety:** for safety of UVGI technicians, fit testing of FFRs Research/Engineering Staff: UVGI system design, technical validation, resources
**Facilities and Electrical:** space allocation, facility construction, resources
**Supply Chain Management:** acquisition of appropriate resources to enable collection
**Facility Leadership:** overall management and priority allocation/determination

Each workgroup member should be positively engaged in the process, individually empowered to pursue the best course of action towards efficient and immediate implementation of workgroup decisions, and allowed to prioritize workgroup tasks by direct action without delay. Communication should be carefully managed, to coordinate individual efforts in smaller teams using direct electronic or voice contacts (better for staff that are not typically active at their desks), and intermittent group updates by email to keep efforts coordinated and progress clear. Facility-wide communication should also be coordinated regarding group progress and implementation plans, and ideally mask collection should commence immediately to allow for immediate decontamination at scale once the workflow is implemented.

## Collection and Labeling Logistics

Our local staff have a strong preference that decontaminated N95 FFRs be returned to the original user, so we developed a procedure for mask collection and tracking based on Nebraska Medicine’s well-established protocol [10]. Additionally, as some pathogens, such as bacterial spores, might survive UV-C irradiation [15], so this “return to original user” approach also prevents potential cross-contamination by these agents. At our facility, masks are labeled on the edges with marker, using both a unique identifier name based on local computer login credentials and the provider’s unit location. Alternative labeling techniques using tags or stickers were not an option given our mask design, as they posed the potential for incomplete mask decontamination due to UV light shading of strap components (which would be re-exposed during mask re-use) or inappropriate stress points on the straps. Although discussed as a hypothetical concern, we are not aware of any data indicating that magic marker writing on the edge of the mask could degrade mask material/function, and we did not observe any instances in which magic marker caused masks to fail fit testing (either before or after decontamination).

We recommended that masks be labeled prior to donning, to minimize the potential for provider/workspace contamination after doffing. We also recommended that care be taken with mask straps to avoid damage, as visible strap stretching from even a single use was occasionally significant. Staff were instructed to avoid wearing cosmetics that could build up on mask surfaces and prevent decontamination. Labeling instructions were disseminated to charge nurses at each unit, posted at collection sites, presented repeatedly at staff meetings, and communicated verbally during visits to collection sites, which improved compliance. However, by the 4^rd^ week after implementation, approximately 20% of masks still remained incompletely labeled, and were likely to be discarded, so this remains an area for improvement.

Proper donning and doffing technique is critical to prevent provider contamination, both immediately after patient care and again when masks are donned, in the event of incomplete decontamination of more resistant pathogens. After doffing each N95, users were instructed to carefully place their masks in brown paper bags, to avoid provider and bag contamination. Each ward placed a mask collection bin with laminated instructions at a central location, and providers used a marker at the collection site to label the bag with name, unit, and date. Our sterile processing service collected masks from each site daily, and these were stored in a central location until ready for decontamination.

In relation to the previously published protocol from Nebraska [10], our collection and labeling protocol is similar, although differs subtly in the following respects: 1. Labeling does not include the date of first use to improve labeling compliance/ease, 2. Brown bags are folded over at the top and collected in closed carts, and 3. Sterile processing staff have been keeping a log sheet of all returned masks (listed by user/unit). Although tracking of individual masks is not a goal of this effort, it allows for user feedback in terms of mask labeling legibility, and also allows for unit-specific tracking of N95 return rates.

## Workflow

The decontamination process itself involves a uni-directional flow of masks through our custom-built UV decontamination suite, in which masks are brought in sealed bins to the “dirty” work area, decontaminated in the adjacent (but separated) UVGI room, and then transferred to the “clean” workroom for packaging into white paper bags. These bags are labeled with the provider’s unique identifier, unit, date/time of UVGI cycle. Masks are marked after each cycle on the mask edge. HVAC for each work area was adjusted such that the decontamination room itself was a negative pressure space, while the clean workroom maintained positive pressure relative to the other spaces.

We plan to decontaminate and re-use the N95 FFRs up to 10 times, as long as they continue to pass repeated user-performed seal testing, based on prior studies of multiple decontamination and wear cycles [13]. Our facility predominantly has the Halyard 46728 “Duckbill” masks, and although we have yet to re-issue masks after decontamination, we have had anecdotal reports of inadequate mask seal after several don/doff cycles (without decontamination) due to stretched straps. Although mask straps for this design could potentially be replaced [23], these observations emphasize the importance of seal tests for each decontaminated mask, at each re-use cycle.

## Throughput

Our current workflow involves hanging masks by their straps on a line between two light sources. This line is connected to hooks on each end, and tensioned by way of rubber bands. The two tensioned lines between the posts can each currently accommodate 20 masks, for the ~30 minute UVGI cycle. Since loading/unloading of the masks occurs within the UV chamber room, it can only be run approximately once per hour. If needed to increase throughput, we will convert to a cantilevered cart system, with protruding arms supporting the tensioned lines. With this improvement, carts could be loaded with masks outside of the UV chamber, while a UV decontamination cycle is taking place. This could increase throughput by about 50–80% without the addition of new UV decontamination lights. Additionally, the addition of reflective material to surround the UVGI chamber could increase the available UV-G energy delivered to each mask, while also improving the number of incident angles, and thus might further increase throughput by reducing exposure times.

## Storage Considerations

Although there are no official guidelines for storage conditions related to post-decontaminated masks, we recommend that masks be stored in a temperature and humidity controlled environment, at least within the manufacturer’s original framework for new masks, but ideally with humidity < 60%. The space required for storage could be substantial, depending on storage modality and the number of masks, and is best estimated with a small group of bagged masks for empiric assessment.

Considerations relating to storage space for N95 FFRs needing decontamination, or decontaminated masks awaiting redeployment, depend greatly on local conditions. Specifically, as new N95s are preferred over decontaminated masks [16], the deployment of decontaminated N95s would likely be coordinated relative to supply/demand balance. However, due to the existence of non-surgical grade FFRs with different performance and fit characteristics, re-deployment of decontaminated, high quality N95 FFRs might be preferable to the use of either non-surgical N95 FFRs or those obtained from community sources.

## Regulatory Considerations

To our knowledge, there are not currently any FDA-approved UVGI N95 decontamination systems. However, the CDC has advised institutions that they could consider decontamination to continue to support their clinical staff [16], and thus, each facility will need to create its own system to provide final approval to the process, likely involving the generation of detailed SOP and Competency documents, similar to those needed for other sterile processing procedures. We would recommend expediting this process, and or obtaining provisional approval to begin decontamination on a provisional basis immediately while paperwork is being finalized, if needed, to prioritize patient/staff protection while regulatory compliance is being addressed.

## Summary

During the current COVID-19 pandemic, PPE supply chains and demand will likely vary significantly in a temporally and regionally specific manner. This supply-demand balance will impact the need for N95 FFR decontamination and the availability of supplies to accomplish this goal. Herein we have devised a methodology that leveraged local resources and supplies to execute a local robust, data-driven, replicable UVGI-based decontamination process.

## Data Availability

All data will be freely disseminated upon request.

## Acknowledgements

We would like to thank David Kagen and Sahana Misra for convening our decontamination workgroup; Darwin Goodspeed for consistent logistic support and encouragement; the VA Portland Electrical Shop and Facilities Management staff for fantastic design implementation and construction assistance; Sherri Atherton (Infection Control) and Ky Dehlinger (Veterinary Medical Unit) for advice; David Cohen and Archie Bouwer for allocation of research space and equipment; Julie Guichot and Oscar Gonzalez for assistance with fit-testing; and Michele Dollar, Grace Chien and Esther Sung for logistic support. We would also like to thank Amy Herr and Alisha Geldert for helpful discussions and comments on this manuscript, and Peter Schnell for assistance with figure preparation. No external funding was explicitly received in support of this work; but the authors would like to acknowledge support by the Department of Veterans Affairs, Veterans Health Administration, Office of Research and Development, Biomedical Laboratory Research and Development Merit Review Awards I01-BX002949 (ES), I01 CX001562 (MH), I01-BX002547 (SMS), a Department of Defense CDMRP Award W81XWH-18–1–0598 (ES), an NIH NINDS 1R21NS102948 (ES), an NIH R01 AI129976 (MH) and an 1R21AI151079–01 (EK). The contents of this manuscript do not represent the views of the U.S. Department of Veterans Affairs or the United States Government.

## Notes

### Competing Interest Statement

The authors have declared no competing interest.

